# Handwriting patterns in Isolated Rapid Eye Movement Sleep Behaviour Disorder

**DOI:** 10.1101/2025.04.01.25325016

**Authors:** Roberta Torricelli, Jack ES Kenny, Emma Bache, Laura Pérez-Carbonell, Brook FR Huxford, Harneek Chohan, Guy Leschziner, Jane Alty, Andrew J Lees, Anette Schrag, Alastair J Noyce, Cristina Simonet

## Abstract

**Background:** Handwriting changes, such as micrographia, are recognised as an early manifestation of Parkinson’s disease (PD). Whilst isolated rapid eye movement sleep behaviour disorder (iRBD) is strongly associated with future PD diagnosis, changes in handwriting remain under-explored.

**Objective:** To assess the handwriting of people with iRBD and develop a rating scale for detection of early disease clinical hallmarks.

**Methods:** This is a cross-sectional study involving 33 people with polysomnography (PSG)- confirmed iRBD and 29 healthy controls. Participants copied a standard sentence using a pen and paper. A graphologist analysed each handwriting script blindly and designed a scale based on observed abnormal patterns which included: micrographia, sentence slope, hidden tremor, retracing, resting marks, irregular shape, excessive pen pressure, and inconsistent word spacing. Each item was scored 0/1 based on their absence/presence. Separately, three blinded movement disorders experts assessed the scripts based on their global clinical impression as well as using the developed rating scale.

**Results:** People with iRBD were slower to complete the task than controls (76.70s (SD = 30.39) vs 61s (SD = 10.71); p=0.004). Hidden tremor was the most common feature amongst the iRBD group (72.0% vs 34.5%; p=0.005), followed by sentence slope (60% vs 24% p=0.005) and pen pressure (48% vs 14%; p=0.006). Micrographia was equally observed in both groups (iRBD 45.4%, controls 41.4% p=0.801). Classification accuracy of the scale for iRBD was higher than expert global assessment (AUC 0.76 vs AUC 0.62, p = 0.029).

**Conclusions:** Writing speed, tremor, pen pressure and sentence slope are handwriting features that warrant further investigation to define early patterns in people with iRBD.

## Introduction

Rapid Eye Movement (REM) sleep behaviour disorder (RBD) is a parasomnia characterised by the loss of normal muscle atonia during REM sleep, leading to dream enactment behaviours (1). It requires video-polysomnography (v-PSG) to confirm the diagnosis. Longitudinal studies have demonstrated a strong link between isolated RBD (iRBD) and synucleinopathies, such as Parkinson’s disease (PD), Dementia with Lewy Bodies (DLB) or Multiple System Atrophy (MSA), with more than 80% of people with iRBD developing one of these later in life (2–4).

In individuals with iRBD, subtle motor signs seem to be the strongest predictor of future parkinsonism; in a large multicenter study including over 1,000 individuals with iRBD, it was found that motor impairment had the highest hazard ratio among 21 potential markers of phenoconversion (2). Additionally, a smaller study suggested that motor dysfunction may precede a formal diagnosis by 6 to 9 years (2). Despite that, early recognition of these features remains challenging primarily due to the limited number of scalable, reliable, and replicable motor assessment tools.

Handwriting analysis is a promising approach to fill this gap. As a complex motor-cognitive task, handwriting integrates voluntary and automatic movements requiring a coordinated interplay of fine motor movements, sensory feedback and cognitive processing (5). This makes it a valuable tool for detecting clinical abnormalities in parkinsonian disorders and dementia. In fact, handwriting changes are a frequent and early manifestation of PD often preceding other motor symptoms (6–10). James Parkinson noted in 1817 that writing difficulties could precede walking impairment (11). Amongst these disturbances, micrographia, defined as a progressive reduction – decrement – in handwriting size and speed over time, is the most well-described handwriting pattern in PD. Originally described by Arnol Pick in 1903 and later linked to PD by Froment approximately two decades later, micrographia remains a clinically relevant marker of parkinsonian motor dysfunction (8, 12–14).

Graphology could be highly valuable in this context, as its core objective is to analyse the dynamics of gestures related to drawing and writing (15). By identifying subtle, premorbid handwriting changes that might otherwise go unnoticed by the untrained eye, graphology offers a unique tool for early detection. Several studies have investigated handwriting features in movement disorders, using either traditional graphological approaches or computational techniques (16–18). Moreover, the benefits of this approach extend beyond letters and sentences to include drawings such as the Archimedean spiral and circles, which are all relevant elements of the clinical motor assessments (19–22).

The aim of this study is to describe handwriting patterns of people with iRBD through the help of a graphologist, and to develop a clinically meaningful, replicable, rating scale to help clinicians and researchers to assist in the assessment and early identification of motor dysfunction in iRBD.

## Methods

### Participants

People with iRBD were identified from the Sleep Clinic at Guy’s & St Thomas’ NHS Foundation Trust, UK, and had undergone overnight v-PSG to confirm the diagnosis of iRBD. Healthy controls were recruited from the PREDICT-PD study; a longitudinal web-based study involving over 10,000 people from the UK general population with the main goal to detect PD early (23). A movement disorders specialist (CS) screened for exclusion criteria in both individuals with iRBD and controls for exclusion criteria which included a formal diagnosis of dementia, PD, and/or any other condition that could interfere with the performance of the handwriting exercise, including essential tremor, motor neuron disease, multiple sclerosis and polyneuropathy. People who were not fluent in English and illiterate were excluded. Similarly, people on medications with potential for causing pharmacological parkinsonism were also excluded.

### Clinical Assessment

Participants were examined by CS (unblinded) using the motor section (III) of MDS-UPDRS and asked about motor symptoms using the MDS-UPDRS-II (Motor Aspects of Experiences of Daily Living, M-EDL) (23). Subthreshold Parkinsonism (SP) was used as a motor outcome measure. The definition for SP was based on the MDS Task Force research criteria for prodromal PD (MDS-UPDRS-III >6 points, excluding postural and kinetic tremor). Participants completed the Montreal Cognitive Assessment (MoCA) to detect cognitive impairment.

### Handwriting Assessment

A clinical rater and neurologist (CS) asked participants to copy the sentence ‘*Mary had a little lamb, its fleece was white as snow*’ three times using a pen and plain paper with their dominant hand and timed them whilst doing so. Separately, an expert graphologist (EB), who was blinded to the participants’ diagnoses, reviewed the handwriting scripts of both groups using a mini handheld magnifying lens so that she could see individual strokes. Handwriting features considered to be ‘abnormal’ were identified and selected to create an 8-item scoring scale:

1. Micrographia: a progressive decrease in letter size within a sentence, followed by a brief increase, and then a repeated reduction with each subsequent sentence
2. Sentence slope: gradient or angle of the line of handwriting
3. Tremor: evidence of irregular pen marks caused by involuntary hand movements
4. Retracing: parts of the handwriting that have been covered by a secondary pen mark
5. Resting marks: ink marks made on the paper by temporary cessation of hand movement
6. Excessive pen pressure: evidence of strong pen pressure applied to the paper creating indentations or excessive ink flow
7. Irregular shape: shape of letters that go against usual formation taught as copy book and variation of letter shape within one handwriting sample
8. Word spacing: space left between handwritten words on a page
9. Handwriting speed: graphologist’s evaluation of the speed and spontaneity of the handwriting

The graphologist scored as 0 or 1 all the items based on the absence or presence of the feature respectively.

Three movement disorders expert neurologists (JA, AJN, AJL), who were blinded to clinical information, independently reviewed scanned copies of the scripts. They were asked to use the handwriting scale developed by the graphologist, excluding the assessment of excessive pen pressure, which could not be evaluated in the scanned versions. They were also asked to classify each script as either abnormal (RBD) or normal (control) based on their overall clinical impression, using their individual clinical judgement.

### Statistical Analysis

Normality of the distribution of data was assessed using D’Agostino K2 test. Mean and standard deviation (SD) were calculated for normally distributed data, whilst interquartile ranges (IQRs) were used for non-normally distributed data. Categorical variables were presented by absolute frequency and percentage, calculating statistical significance using Fisher’s exact test, whilst quantitative data for demographic and motor outcomes were compared using Welch’s test for unequal variances.

The classification accuracy of the handwriting rating scale was determined using the Wilson/Brown method, including the generation of Receiver Operator Characteristic (ROC) curves to evaluate its performance at different thresholds. A cut-off that maximised Youden’s J index was selected for each variable.

Inter-rater variability of the three movement disorder specialists was calculated using Kappa index.

All statistical tests were two-tailed. Bonferroni correction method was used to adjust the cut-off for evidence of association in the handwriting scale, which included nine variables, setting the p-value at <0.006 (0.05/9). Data analysis was carried out using STATA v.13 (StataCorp, College Station, TX).

## Results

### Demographics and Comorbidities

A total of 34 people with iRBD and 29 controls were recruited. One participant with iRBD fulfilled the diagnosis of PD upon clinical examination, so their data was subsequently excluded from the analysis. In the final sample, there were 33 participants with iRBDs and 29 controls. Mean disease duration from symptoms onset in people with iRBD was 10.6 years (SD 6.87), with a delay in the RBD diagnosis since noticeable symptom onset of 3 years (IQR 1 to 3). People with iRBD were younger than controls (iRBD mean age (SD): 68.9 years (8.1)) vs controls: 74.9 years (5.4), p<0.001) but the percentage of male participants was similar (controls: 23/29 (79.3%) vs iRBD: 30/33 (90.9%); p=0.283). There were no group differences in MoCA scores (iRBD mean (SD) 26.39 (2.96), controls 27.45 (1.86), p=0.095) or years of education (iRBD mean (SD) 19.5 (1.1), controls 20.7 (0.6), p=0.313) (see Table 1).

**Table 1:** Clinical information and handwriting features.

|  |  | *iRBD* | *Controls* | *P-values* |
| --- | --- | --- | --- | --- |
| *Participants* | n | 33 | 29 |  |
| *Age (yrs)* | Mean (SD) | 68.9 (8.1) | 74.9 (5.4) | <0.001* |
| *Males* | n (%) | 30 (90.9) | 23 (79.3) | 0.283 |
| *Education (yrs)* | Mean (SD) | 19.4 (1.1) | 20.7 (0.6) | 0.313 |
| *Disease duration (yrs)* | Mean (SD) | 10.6 (6.87) | - |  |
| *Diagnostic Delay (yrs)* | Median (IQR) | 3 (1-3) | - |  |
| *MoCA score* | Mean (SD) | 26.39 (2.96) | 27.45 (1.86) | 0.095 |
| *Handwriting Features* |  |  |  |  |
| 1. *Micrographia* | n (%) | 15 (45.40) | 12 (41.38) | 0.801 |
| 2. *Sentence slope* | n (%) | 20 (60.60) | 7 (24.14) | 0.005* |
| 3. *Tremor* | n (%) | 24 (72.73) | 10 (34.48) | 0.005* |
| 4. *Retracing* | n (%) | 9 (27.27) | 4 (13.79) | 0.227 |
| 5. *Resting marks* | n (%) | 4 (12.12) | 2 (6.90) | 0.676 |
| 6. *Excessive pen pressure* | n (%) | 16 (48.48) | 4 (13.79) | 0.006* |
| 7. *Irregular shape* | n (%) | 6 (18.18) | 0 | 0.026* |
| 8. *Word spacing* | n (%) | 9 (27.27) | 3 (10.34) | 0.116 |
| 9. *Handwriting speed* | Mean (SD) | 76.70 (30.39) | 61 (10.71) | 0.004* |
| *Handwriting scale score* | Mean (SD) | 3.18 (1.44) | 1.69 (1.58) | <0.001* |

Handwriting scale based on the presence (score 1) or absence (score 0): overall score includes items 1-8; Handwriting speed: handwriting task duration (including the three sentences); “\*” denotes statistical significance defined as $P < 0.05$ . 1) Welch’s test for unequal variances. 2) Fisher’s exact test

### Motor Assessment

Eleven people with iRBD (33.3%) fulfilled criteria for SP. In contrast, only 1 healthy control scored 7 points in the MDS-UPDRS-III (excluding action and postural tremor). On average, the iRBD group scored 4 points higher than controls on the MDS-UPDRS-III (mean (SD): 7.24 (4.81) vs. 3.0 (2.25); p<0.001). Additionally, people with iRBD were more prone to report motor symptoms than controls as reflected in their higher MDS-UPDRS-II total scores (mean (SD): 2.51 (3.16) vs. 1.41 (1.68); p=0.04).

### Handwriting Assessment

The iRBD group took an average of 15 seconds longer to write three sentences than controls (mean time (SD); iRBD: 76.70 seconds (30.39) vs control 61 seconds (10.71); p=0.004). In fact, handwriting speed was found to be able to correctly detect iRBD participants from controls with 63.6% sensitivity and 75.9% specificity when using a cut-off of 21 seconds (AUC 0.71; 95% CI, 0.58 to 0.84). By increasing the cut-off 24 seconds, handwriting speed improved its accuracy to detect iRBD with SP from those without SP, achieving a detection rate of 81.8% for 82.8% specificity (AUC 0.86; 95% CI, 0.68 to 1.00).

Several handwriting differences were found by the graphologist amongst the iRBD group (Table 1). Markers of tremor were the most common feature amongst the iRBD group (present in 72.0% vs 34.5% respectively; p=0.005), followed by sentence slope (60% vs 24%; p=0.005) and increased pen pressure (48% vs 14%; p=0.006). It was noteworthy that, in 10 iRBD participants and in 6 controls, tremor was only visible in handwriting scripts and not with the naked eye during MDS-UPDRS assessment. Examples of handwriting scripts considered abnormal according to graphologist impression criteria are presented in Figure 1 and Figure 2. Despite the above findings, the proportion of people with micrographia in the iRBD group did not differ from controls (15 people with iRBD (45.4%) and 12 controls (41.4%); p=0.801).

**Figure 1:**
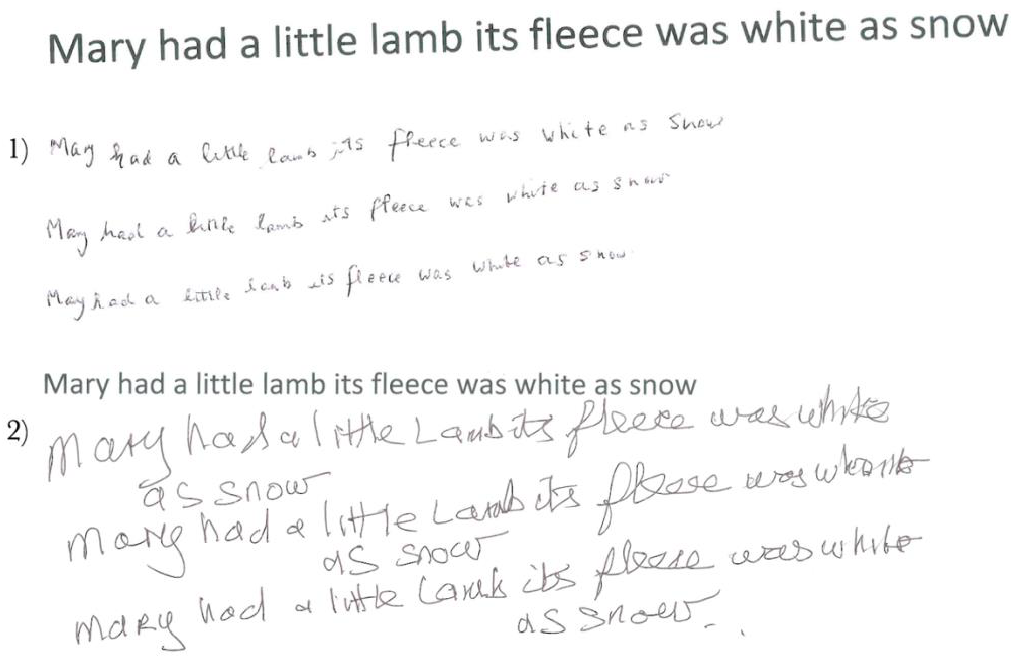
Examples of Abnormal Handwriting. 1) Individual with established PD and micrographia: progressive reduction in handwriting size across sentences (e.g. ‘white’ and ‘snow’). 2) Participant with iRBD and dysgraphia: untidy (irregular letter and word shape). Note sentence slope in both examples.

**Figure 2:**
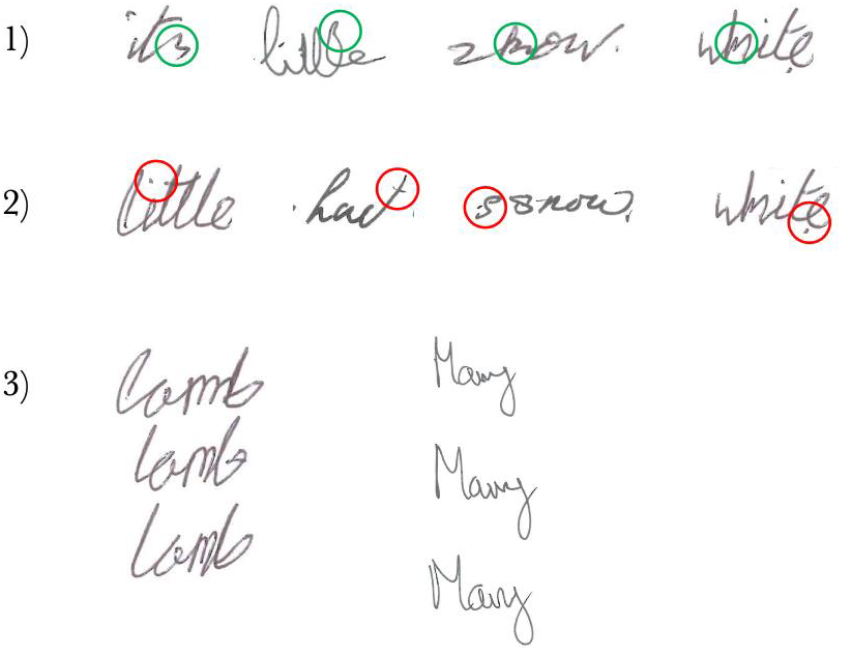
Examples of Handwriting Markers in iRBD. 1) hidden tremor in green, 2) resting marks in red and 3) irregular word shape across sentences.

Specificity and sensitivity were calculated to evaluate the accuracy of the scale; indeed, we found that when administered by a graphologist, it was able to correctly detect iRBD with 75.8% specificity and 68.9% sensitivity (cut-off: four points; AUC 0.77; 95% CI, 0.64 to 0.89). However, it was less accurate in identifying iRBD with SP from iRBD without SP with a detection rate of 72.4% for 69.7% sensitivity (cut-off: three points; AUC 0.81; 95% CI, 0.60 to 0.88) compared to handwriting speed.

We found a high inter-observer variability (Kappa index −0.08) amongst the 3 movement disorders expert neurologists when asked to provide their global impression and score the items in the handwriting scale. For that reason, we decided to use the best-performing expert assessment, as defined by the expert with the lowest standard error and highest sensitivity, to be compared with the graphologist assessment and found that the scale was more accurate (AUC 0.62 vs AUC 0.77, p=0.029).

In terms of the MDS-UPDRS-III, it had a similar accuracy to the handwriting scale when implemented by a graphologist to correctly detect iRBD (cut-off: three points; AUC 0.81; 95% CI, 0.68 to 0.87; 69.7% sensitivity and 72.4% specificity).

## Discussion

To our knowledge, this is the first study to explore handwriting features in people with iRBD. We developed a simple handwriting scale that can be applied in practice, without the need for sophisticated equipment, allowing for the identification of distinctive handwriting patterns in iRBD beyond micrographia. In addition to reduced writing speed, we observed distinctive features in iRBD including altered sentence slope, hidden tremor within letters, irregular letter shape, and increased pen pressure. The fact that our scale, applied by a blinded graphologist, showed similar accuracy to the MDS-UPDRS-III, administered by an unblinded clinical rater, further strengthens the robustness of our results.

Micrographia has been linked to dopamine depletion within cortico-subcortical networks, and it is used as a supportive diagnostic feature to test for PD medication response (25). However, it can also be used as a differential diagnostic tool to distinguish between PD and atypical parkinsonism, such as in Progressive Supranuclear Palsy (PSP) which, in contrast to PD, classically presents with consistent small handwriting without decrement (10, 26). Interestingly, micrographia did not discriminate iRBD in this cohort suggesting that additional handwriting parameters related to movement execution beyond letter size (dysgraphia) may emerge earlier in the early disease course.

Dysgraphia, defined as a disorder of mechanical handwriting skills, has been proposed as a broader term encompassing handwriting impairments in PD, which share similarities with developmental dysgraphia in children. To more comprehensively capture the spectrum of handwriting disturbances in PD, some authors advocate using the term “dysgraphia” instead of restricting analysis exclusively to letter size (27). This broader conceptualisation might facilitate the identification of handwriting abnormalities beyond micrographia, including spatial, temporal, and pressure-related disturbances.

Handwriting and drawing analysis have also been used in the dementia field (28, 29) which has important implications for iRBD, given that a proportion of individuals with this condition will develop DLB. Graphology is traditionally defined as the analysis of an individual’s psychological structure through their handwriting and has been applied in various fields where critical decision-making is essential, such as forensic evidence evaluation, criminology, and mental health disease analysis – especially to historical texts (30, 31, 32, 33). Incorporating graphology expertise into handwriting assessments helped identifying distinct handwriting patterns in iRBD. Follow-up of these participants will be crucial to know who will follow a predominantly cognitive (DLB) or motor (PD) disease trajectory.

Several limitations should be considered in this study. An important one, is the difference in age between the iRBD and the control groups used in the handwriting analysis, with the former being younger. Of note, the older mean age of the control group should count against, not for, a confounding effect on the handwriting assessment scores of participants, as age itself is associated with a further decline in motor function in both iRBD and PD (34–36). Educational background may also represent a confounding factor in handwriting analysis. To minimise this, we chose a sentence that it is commonly learned in school. However, both groups were comparable in terms of years of education. We did not account for other factors that might have an impact on our results such as writing literacy (time used to writing each day), pain/arthritis in hands, anxiety and fatigue.

Another limitation is the absence of a second graphologist to explore whether there also exists interrater variability among handwriting experts, similar to what has been observed among movement disorders specialists. Reasons for their interrater variability could be the lack of consensus on specific handwriting features associated with iRBD. Additionally, the quality of scanned handwriting samples may differ from the original documents, potentially making handwriting analysis more challenging for movement disorder experts compared to the graphologist. Movement disorder specialists were further disadvantaged in their assessments due to their inability to evaluate pen pressure or time to complete the task in the scanned copies, which was one of the most distinctive handwriting features in iRBD and could have negatively impacted their overall performance when rating the handwriting samples. As with the MDS-UPDRS, training may be necessary to improve the accuracy and reliability of handwriting assessments based on this scale.

Moreover, although having a reference sentence is necessary to standardise assessment, research has shown that handwriting abnormalities, in particular micrographia, become more evident during free writing and could instead be masked by copying tasks (37). This could explain why micrographia did not differ between the groups. Nonetheless, the lack of visual cues on the paper, such as lines or squares, would still have allowed for more of the handwriting abnormalities and potential sentence inclination to emerge (38).

It is important to acknowledge that our results might not be generalisable to other cohorts due to our small sample size from a predominantly White UK-based population. Additionally, there might be a healthy volunteer bias as individuals who participate in research studies are often healthier than those recruited from healthcare or community settings (39)

Lastly, the cross-sectional nature of our study limits our ability to assess disease progression and determine whether specific handwriting patterns may serve as early markers of neurodegenerative disease. Longitudinal follow-up studies would be essential to validate this handwriting scale and establish its predictive value in identifying individuals at risk for PD.

To conclude, we present handwriting signatures in iRBD which could be capable of detecting early motor and cognitive manifestations. However, our findings remain exploratory and warrant further investigation towards the development of tools capable of detecting handwriting signatures in iRBD.

## Data Availability

All data produced in the present study are available upon reasonable request to the authors

## Acknowledgments

There are no acknowledgements.

## Author Roles

1. Research project: A. Conception, B. Organization, C. Execution;
2. Statistical Analysis: A. Design, B. Execution, C. Review and Critique;
3. Manuscript Preparation: A. Writing of the first draft, B. Review and Critique.

RT: 1A, 2C, 3A

JESK: 1A, 2C, 3A

EB: 1C

LPC: 1B, 2C, 3B

BFRH: 1B

HC: 1B

GL: 1B, 2C, 3B

JA: 1C, 3C

AJL: 1A, 1C, 3B

AS: 1C, 2C, 3B

AJN: 1C, 2C, 3B

CS: 1A, 1B, 1C, 2A, 2B, 3B

## Ethical Compliance Statement

Ethics approval was granted by the Queen Square Research Ethics Committee (09/H0716/48). Participants received verbal and written information about the study and appropriately consented.

